# Hospital Admission Rates, Length of Stay and In-hospital Mortality for Common Acute Care Conditions in COVID-19 vs. Pre-COVID-19 Era

**DOI:** 10.1101/2020.08.04.20167890

**Authors:** Adeel A. Butt, Anand B. Kartha, Naseer A. Masoodi, Aftab Mohammad Azad, Nidal Ahmad Asaad, Mohamad Ussama Alhomsi, Huda A. H. Saleh, Roberto Bertollini, Abdul-Badi Abou-Samra

## Abstract

**Background:** Impact of COVID-19 upon acute care admission rates and patterns are unknown. We sought to determine the change in rates and types of admissions to tertiary and specialty care hospitals in the COVID-19 era compared with pre-COVID-19 era.

**Methods:** Acute care admissions to the largest tertiary care referral hospital, designated national referral centers for cardiac, cancer and maternity hospital in the State of Qatar during March 2020 (COVID-19 era) and January 2020 and March 2019 (pre-COVID-19 era) were compared. We calculated total admissions, and admissions for eight specific acute care conditions, in-hospital mortality rate and length of stay at each hospital.

**Results:** A total of 18,889 hospital admissions were recorded. A sharp decline ranging from 9%-75% was observed in overall admissions. A decline in both elective and non-elective surgeries was observed. A decline of 9%-58% was observed in admissions for acute appendicitis, acute coronary syndrome, stroke, bone fractures, cancer and live births, while an increase in admissions due to respiratory tract infections was observed. Overall length of stay was shorter in the COVID-19 period possibly suggesting lesser overall disease severity, with no significant change in in-hospital mortality. Unadjusted mortality rate for Qatar showed marginal increase in the COVID-19 period.

**Conclusions:** We observed a sharp decline in acute care hospital admissions, with a significant decline in admissions due to seven out of eight acute care conditions. This decline was associated with a shorter length of stay, but not associated with a change in in-hospital mortality rate.

## Introduction

The ongoing COVID-19 pandemic has strained global healthcare capacity to the breaking point in many countries.[1-4] Various approaches have been employed by different countries in response to the surge of cases, including physical distancing measures, targeted lockdowns, wider movement restrictions, travel bans, and closing of places of mass gatherings. Despite these measures, the number of persons with COVID-19 infection requiring hospitalization has exceeded acute and critical care bed capacity. To address acute and critical care bed capacity issues, most countries canceled or postponed elective surgeries and other admissions deemed non-vital to short-term patient outcomes. Early anecdotal reports in mainstream media noted a decrease in hospital admissions due to “heart attacks, strokes and even appendicitis”.[5] This was confirmed by more recent reports noting a decline in admissions due to acute coronary syndrome, a decline in ST-segment elevation cardiac catheterization laboratory activations, a decrease in stroke imaging procedure performed, and an increase in out of hospital cardiac arrest.[6-9] An increase in emergency medical services arrival time and a decrease in by-stander initiated cardiopulmonary resuscitation was also noted.[7] The effect of COVID-19 pandemic upon admissions for general medical and surgical care, oncology services and obstetric services, and the effect of any change on in-hospital and overall community mortality rates is not known. Our aim was to determine the change in rates and reasons for hospitalization at four major referral centers in the State of Qatar. We chose the country’s largest and main tertiary care hospital, and dedicated hospitals and national referral centers for comprehensive cardiovascular care, cancer care and obstetric and gynecologic services.

## Methods

The study was conducted at Hamad Medical Corporation (HMC), the State owned and operated healthcare system, which provides approximately 85% of acute care inpatient bed capacity in Qatar. The flagship and the largest hospital is Hamad General Hospital (HGH), which is the tertiary care referral center located in the capital city, Doha. Comprehensive cardiac care is provided at the Heart Hospital, which is the main designated facility in the country for primary percutaneous interventions for acute coronary syndrome. Cancer care is provided at the National Center for Cancer Care and Research (NCCCR), which is the only facility in the country providing comprehensive cancer care services, including chemotherapy, radiation therapy, and cyberknife services. Women’s Wellness and Research Center (WWRC) is the largest provider of obstetric and gynecologic care in the country. These facilities were chosen since they represent the spectrum of acute tertiary and specialty care in the country. All hospitals use the same electronic health records system (Cerner®, Kansas City, MO, USA) which is interconnected, with patients retaining the same unique health identification number across all hospitals. All hospitals are accredited by The Joint Commission International and the clinical laboratories are accredited by the College of American Pathologists. All hospital discharges are reviewed by expert coders to assign the primary and secondary reasons for hospital admission. Up to ten reasons are recorded at the time of discharge using International Classification of Diseases, 10^th^ edition Australian Modification (ICD-10-AM).

The first case of COVID-19 in Qatar was diagnosed on February 28, 2020. As a matter of national policy, these four hospitals were designated to be COVID-19 free facilities. Any patient with known COVID-19 infection was admitted to a separate COVID-19 facility. Patients diagnosed incidentally or upon routine testing for COVID-19 after admission to any of these hospitals were evaluated and transferred to a designated COVID-19 facility, unless they were medically unstable for transfer. We determined the number of hospital discharges at each of the four hospitals for the month of March 2020 and compared it with January 2020 and March 2019. We omitted February 2020 since changes in hospital flow were being implemented in anticipation of the first wave of COVID-19 patients. All inpatient admissions at the four hospitals with a length of stay of more than one day were included. Data retrieved included patient demographics (age, sex and nationality), admission and discharge dates and times, principal and up to nine additional discharge diagnoses, mode of arrival at the hospital, disposition, and surgical procedures performed (elective vs. nonelective).

We tabulated the overall number of admissions for each hospital across the three study periods, as well as the number of elective and non-elective surgical procedures performed, average daily admissions, average length of stay and in-hospital mortality. Hospital utilization metrics, including mode of presentation to the hospital for that particular episode, and disposition were tabulated for each hospital, by study period.

Through consensus among co-authors, eight common diagnoses/conditions were chosen to represent the spectrum of conditions that generally require acute hospital care. These included acute appendicitis, acute coronary syndrome, other cardiovascular disease diagnoses (including cardiac arrhythmias, congestive heart failure and angina pectoris without acute myocardial infarction), stroke, acute bone fractures, cancers, live births, and respiratory tract infections (excluding tuberculosis). Two study team members independently reviewed all diagnoses and identified and assigned them to one of the categories listed above. These diagnostic groups were tabulated by study period, and the percentage change in each category was calculated.

To explore any correlation between acute care admission rates, in-hospital mortality and overall mortality rate in the State of Qatar, we retrieved publicly available data published by the Planning and Statistics Authority, State of Qatar, which publishes monthly reports on the total population and other vital statistics for persons residing in Qatar for that month. Number of deaths and the total population of Qatar for the months included in the study were retrieved to calculate the unadjusted death rate for each month.

## Ethical Approval

The study was reviewed and approved by the Institutional Review Board at Hamad Medical Corporation (MRC-05-034).

## Results

A total of 18,889 hospital admissions were recorded for the three study periods. **(Table 1)** There were 7,545 admissions at HGH, 794 at HH, 396 at NCCCR and 10,154 at the WWRC. Compared with January 2020, there was a 28.0% reduction in admissions at HGH, 59.6% reduction at HH, 45.7% reduction at NCCCR and a 14.7% reduction at the WWRC. Compared with the same month in 2019, the reduction was 8.6% at HGH, 51.8% at HH, 75.5% at NCCCR and a 21.5% at the WWRC. Number of elective surgeries dropped by 6.8% at the WWRC to 68.6% at HGH from January 2020 to March 2020, while non-elective surgeries showed a smaller decline. Average daily admissions and average length of stay also declined at all facilities over both time period comparisons. **(Table 1)** In-hospital mortality rate was numerically lower at all hospitals for all time period comparisons except at HGH between March 2010 and January 2020. However, these comparisons were not statistically significant. There were no significant changes in terms of mode of presentation to the hospital or disposition at any of the facilities. **(Table 2)**

**Table 1.** Change in number of total admissions, surgeries, daily admissions, length of stay and in-hospital mortality. (T0 = March 2020; T1 = January 2020; T2 = March 2019)

|  | Hamad General Hospital | Heart Hospital | National Center for Cancer Care and Research | Women's Wellness and Research Center |
| --- | --- | --- | --- | --- |
| **Total admissions** |  |  |  |  |
| COVID-19 period (T0) | 2,242 | 193 | 94 | 3,020 |
| Pre-COVID-19 period (T1) | 2,869 | 308 | 137 | 3,465 |
| Same period previous year (T2) | 2,434 | 293 | 165 | 3,669 |
| % Change T0 – T1 | -28.0% | -59.6% | -45.7% | -14.7% |
| % Change T0 – T2 | -8.6% | -51.8% | -75.5% | -21.5% |
| **Elective Surgeries** |  |  |  |  |
| COVID-19 period (T0) | 312 | 98 | 10 | 237 |
| Pre-COVID-19 period (T1) | 526 | 157 | 11 | 253 |
| Same period previous year (T2) | 473 | 151 | 9 | 288 |
| % Change T0 – T1 | -68.6%* | -60.2%<sup>†</sup> | -10.0%<sup>†</sup> | -6.8%<sup>†</sup> |
| % Change T0 – T2 | -51.6%* | -54.1%<sup>†</sup> | 10.0%<sup>†</sup> | -21.5%<sup>†</sup> |
| **Non-elective Surgeries** |  |  |  |  |
| COVID-19 period (T0) | 548 | 59 | 1 | 391 |
| Pre-COVID-19 period (T1) | 626 | 81 | 3 | 392 |
| Same period previous year (T2) | 661 | 68 | 1 | 442 |
| % Change T0 – T1 | -14.2%* | -37.3%<sup>†</sup> | -200.0%<sup>†</sup> | -0.3%<sup>†</sup> |
| % Change T0 – T2 | -20.6%* | -15.3%<sup>†</sup> | 0.0%<sup>†</sup> | -13.0%<sup>†</sup> |
| **Average daily admissions** |  |  |  |  |
| COVID-19 period (T0) | 72.32 | 6.23 | 3.03 | 97.42 |
| Pre-COVID-19 period (T1) | 92.55 | 9.94 | 4.42 | 111.77 |
| Same period previous year (T2) | 78.52 | 9.45 | 5.32 | 118.35 |
| Change T0 – T1 | -20.23 | -3.71 | -1.39 | -14.35 |
| Change T0 – T2 | -6.2 | -3.22 | -2.29 | -20.93 |
| **Average length of stay** |  |  |  |  |
| COVID-19 period (T0) | 3.97 | 6.84 | 5.65 | 2.24 |
| Pre-COVID-19 period (T1) | 5.81 | 9.97 | 13.44 | 2.93 |
| Same period previous year (T2) | 6.68 | 12.64 | 13.38 | 3.13 |
| Change T0 – T1 | -1.84* | -3.13* | -7.79* | -0.69* |
| Change T0 – T2 | -2.71* | -5.8* | -7.73* | -0.89* |
| **In-hospital mortality** |  |  |  |  |
| COVID-19 period (T0) | 1.16% | 1.04% | 4.26% | 0.13% |
| Pre-COVID-19 period (T1) | 0.98% | 2.27% | 8.76% | 0.20% |
| Same period previous year (T2) | 1.52% | 2.39% | 8.48% | 0.14% |
| Change T0 – T1 | 0.18%<sup>†</sup> | -1.23%<sup>†</sup> | -4.50%<sup>†</sup> | -0.07%<sup>†</sup> |
| Change T0 – T2 | -0.36%<sup>†</sup> | -1.35%<sup>†</sup> | -4.22%<sup>†</sup> | -0.01%<sup>†</sup> |

\* P≤0.01
<sup>†</sup> P>0.05

**Table 2.** Utilization metric at four hospitals during the study periods.

|  | March 2020<br>(T0) | January 2020<br>(T1) | March 2019<br>(T2) |
| --- | --- | --- | --- |
| **Hamad General Hospital** |  |  |  |
| Number of total admissions, N | 2,242 | 2,869 | 2,434 |
| Admission and flow metrics |  |  |  |
| Self-presentation to ED | 71.10% | 71.91% | 70.05% |
| Direct admission from clinic | 20.61% | 18.51% | 21.94% |
| Transfer from another hospital | 0.13% | 0.07% | 0.16% |
| Other including via ambulance | 8.16% | 9.52% | 7.85% |
| Disposition |  |  |  |
| Home | 92.60% | 94.11% | 93.14% |
| Another facility | 2.85% | 0.91% | 0.74% |
| Died | 1.16% | 0.98% | 1.52% |
| Other | 3.39% | 4.01% | 4.60% |
| **Heart Hospital** |  |  |  |
| Number of total admissions, N | 193 | 308 | 293 |
| Admission and flow metrics |  |  |  |
| Self-presentation to ED | 63.21% | 58.44% | 61.43% |
| Direct admission from clinic | 12.95% | 14.94% | 11.60% |
| Transfer from another hospital | 3.63% | 5.52% | 3.41% |
| Other including via ambulance | 20.21% | 21.10% | 23.55% |
| Disposition |  |  |  |
| Home | 96.89% | 94.48% | 94.20% |
| Another facility | 0% | 0% | 0% |
| Died | 1.04% | 2.27% | 2.39% |
| Other | 2.07% | 3.25% | 3.41% |
| **National Center for Cancer Care and Research** |  |  |  |
| Number of total admissions, N | 94 | 137 | 165 |
| Admission and flow metrics |  |  |  |
| Self-presentation to ED | 45.74% | 46.72% | 39.39% |
| Direct admission from clinic | 40.43% | 32.85% | 37.58% |
| Transfer from another hospital | 0.00% | 0.73% | 0.61% |
| Other including via ambulance | 13.83% | 19.71% | 22.42% |
| Disposition |  |  |  |
| Home | 92.55% | 86.13% | 89.09% |
| Another facility | 1.06% | 1.46% | 0.61% |
| Died | 4.26% | 8.76% | 8.48% |
| Other | 2.13% | 3.65% | 1.82% |
| **Women's Wellness and Research Center** |  |  |  |
| Number of total admissions, N | 3,020 | 3,465 | 3,669 |
| Admission and flow metrics |  |  |  |
| Self-presentation to ED | 39.37% | 40.61% | 40.45% |
| Direct admission from clinic | 16.52% | 14.66% | 17.23% |
| Transfer from another hospital | 0.07% | 0.17% | 0.14% |
| Other including via ambulance | 44.04% | 44.56% | 42.19% |
| Disposition |  |  |  |
| Home | 99.21% | 98.96% | 99.24% |
| Another facility | 0.26% | 0.40% | 0.22% |
| Died | 0.13% | 0.2% | 0.14% |
| Other | 0.40% | 0.43% | 0.41% |

Between January 2020 and March 2020, there was a decrease in admissions for seven of the eight conditions. **(Table 3)** Admissions were lower for acute appendicitis (−17.4%), acute coronary syndrome (−57.9%), other cardiovascular disease diagnoses (−48.4%), stroke (−30%), bone fractures (−8.7%), cancer (−12.9%), and live births (−8.8%), while admissions were higher for respiratory tract infections (+15.1%). Respiratory tract infection admissions included 88 patients with a diagnosis of “coronavirus infection” who were diagnosed upon routine testing after admission. Between March 2019 and March 2020, there was also a decrease in admissions for the same seven of the eight conditions. **(Table 3)** Admissions were lower for acute appendicitis (−22.8%), acute coronary syndrome (−50.0%), other cardiovascular disease diagnoses (−81.3%), stroke (−31.3%), bone fractures (−38.8%), cancer (−23.9%), and live births (−17.0%), while admissions were higher for respiratory tract infections (+56.1%). **(Table 3)**

**Table 3.** Change in number of patients with selected diagnoses across four hospitals during the study periods.

|  | **March 2020**<br>**(T0)** | **January 2020**<br>**(T1)** | **March 2019**<br>**(T2)** | **T0 – T1**<br>**%** | **T0 – T2**<br>**%** |
| --- | --- | --- | --- | --- | --- |
| Acute appendicitis | 92 | 108 | 113 | -17.4% | -22.8% |
| Acute coronary syndrome | 114 | 180 | 171 | -57.9% | -50.0% |
| Other cardiovascular disease diagnoses | 64 | 95 | 116 | -48.4% | -81.3% |
| Stroke | 83 | 108 | 109 | -30.1% | -31.3% |
| Bone fracture(s) | 183 | 199 | 254 | -8.7% | -38.8% |
| Cancer | 155 | 175 | 192 | -12.9% | -23.9% |
| Live births | 1,291 | 1,405 | 1,510 | -8.8% | -17.0% |
| Respiratory tract infections<sup>1</sup> | 337<sup>2</sup> | 286 | 148 | 15.1% | 56.1% |

<sup>1</sup> excluding tuberculosis;
<sup>2</sup> includes 88 cases of coronavirus disease

Unadjusted mortality rate (number of deaths recorded in a given month divided by the population in the same month) per 100,000 population was 7.91 for March 2020, 6.92 for January 2020, and 6.48 for March 2019. Total number of deaths recorded were 221 for March 2020, 192 for January 2010, and 179 for March 2019. The difference in number of deaths was not statistically significant for comparison between March 2020 vs. January 2020 (p=0.2), but was significant for the comparison between March 2020 vs. March 2019 (p=0.05). **(Table 4)**

**Table 4.** Population statistics for the State of Qatar for the study periods.

|  | **March 2020**<br>**(T0)** | **January 2020**<br>**(T1)** | **March 2019**<br>**(T2)** | **T0 - T1**<br>**%** | **T0 - T2**<br>**%** |
| --- | --- | --- | --- | --- | --- |
| Total population | 2,795,484 | 2,773,221 | 2,760,586 | 0.8 | 1.2 |
| Total deaths | 221 | 192 | 179 | 13.1* | 19.0<sup>†</sup> |
| Unadjusted death rate | 7.91 | 6.92 | 6.48 | 12.5 | 18.1 |

\* P=0.2
<sup>†</sup> P=0.05

## Discussion

To our knowledge, this is the first study to quantify the change in admission volumes and reasons for admission for common acute care conditions in the COVID-19 vs. pre-COVID-19 era. We observed a sharp decline in all studied conditions except respiratory tract infections, where an increase in admissions was observed.

Many countries and healthcare systems around the world enacted policies to reduce acute care hospital admissions by postponing elective admissions. What may not have been anticipated was a decline in non-elective admissions, which would otherwise be considered essential for optimal outcomes for the patients. While it is possible that some patients with urgent or emergent acute conditions (e.g. acute myocardial infarction, acute appendicitis, stroke, etc.) may survive without being admitted to a hospital, lack of supervised medical care would certainly lead to overall poorer short and long-term outcomes and a higher mortality risk. We could not identify any factor other than COVID-19 pandemic related factors to account for the decline in acute care admissions. There are several possible reasons for decline in both elective and non-elective care admissions. These include patients adhering to the physical distancing recommendations, worries about contracting the infection in a healthcare facility, inability to find transportation to the hospital, critical illness which may have affected the cognitive or physical ability to seek care, and avoiding hospital visits for problems perceived to be less than critical.

Another possible reason may be overly liberal criteria for admitting patients in previous months, some of which may not have been necessary. Expanding primary healthcare services in Qatar may also have prevented some soft cases from being referred for admission to an acute care facility. Further studies are needed to determine the precise reasons for this decline in order to ensure that appropriate and timely care is provided to those who need such care.

Consequences of a decline in acute care admissions can be devastating for those with most severe illnesses requiring supervised medical and surgical care. We observed a reduction in admissions for multiple acute and potentially critical conditions. If patients with severe or critical illness present less frequently to acute care facilities, it is conceivable that the overall case mix index for admitted patients would be lower and reflect in terms of shorter length of stay and lower in-hospital mortality. We did indeed observe a significantly shorter length of stay across all hospitals included in the study. While in-hospital mortality trend was observed to be numerically lower for most comparisons, none of it reached statistical significance likely due to small number of in-hospital deaths. A concomitant trend towards a higher overall unadjusted death rate in the country was observed, though this must be interpreted with extreme cautions since we only studied three months data and did not ascertain the long-term trends or the reasons for this variation. This is an important hypothesis-generating observation and must not be interpreted as a causal link. The overall crude mortality rate in the country is particularly low reflecting the demographic pattern and the young population of Qatar

The increase in admissions due to respiratory tract infections is likely a reflection of heightened awareness of COVID-19 infection and increased vigilance practiced by healthcare practitioners. We did observe an increase in number of patients who were admitted under isolation precautions in the March 2020, which supports this impression. (data not shown) The four hospitals included in this study were designated to be COVID-19 free facilities, and all diagnosed patients were admitted or transferred to designated facilities for COVID-19 patients. The number of admissions due to respiratory tract infections may have been higher if COVID-19 patients had free access to these hospitals. This policy likely kept the nosocomial COVID-19 infection rate and infection among healthcare workers at a very low level in these facilities. Cohorting of patients also led to more efficient use of resources, both in terms of healthcare personnel and equipment.

Our study raises important questions about future pandemic planning. Follow up studies are needed to determine whether this observed decline in admissions will translate to more severe presentations, late diagnosis, or long-term disability related to these diagnoses. For example, determining the proportion of patients presenting with ruptured appendix, more advanced cancer, more residual weakness or disability after stroke, and higher rates of advanced heart failure can lead to policies targeting such patients in any future pandemic setting, and devising strategies to get them in appropriate care setting earlier. It is also important to understand the magnitude and burden of these consequences when current travel and physical distancing restrictions are removed. Knowing this burden will be critical in planning for the possible surge of patients once these restrictions are lifted.

The strengths of our study include evaluating multiple hospitals which see a variety of acute care conditions and are national referral centers. A variety of acute care conditions were studied to provide a broad understanding of the change in admission patterns. We also studied national mortality trends in an attempt to understand large scale consequences of our findings. Despite numerous strengths, these data need to be interpreted with caution, as they represent only a snapshot in time. Particularly, any correlation with national mortality statistics must be interpreted with extreme caution since numerous factors may affect those statistics. No causal inference can be drawn from the current study regarding the impact of our findings on overall mortality rates for the entire country.

In summary, we observed a significant decline in hospital admissions across several hospitals in a national healthcare system, with a significant decline in admissions due to seven out of eight acute care conditions studied. This decline was associated with a shorter length of stay, but not associated with a change in in-hospital mortality rate. A possible small increase in unadjusted mortality rate in the country requires further study to determine if there is any correlation with the change in hospital admission rates.

## Data Availability

Data is not publicly available

## Notes

### Competing Interest Statement

The authors have declared no competing interest.

### Clinical Trial

N/A

### Funding Statement

This study was funded by the Medical Research Center, Hamad Medical Corporation, Doha, Qatar. (PI: Prof. Adeel A. Butt)

